# Prognostic factors for mortality, ICU and hospital admission due to SARS-CoV-2: A systematic review and meta-analysis of cohort studies in Europe

**DOI:** 10.1101/2022.03.24.22272870

**Authors:** Constantine I. Vardavas, Alexander G. Mathioudakis, Katerina Nikitara, Kimon Stamatelopoulos, Georgios Georgiopoulos, Revati Phalkey, Jo Leonardi-Bee, Esteve Fernandez, Dolors Carnicer-Pont, Jørgen Vestbo, Jan C. Semenza, Charlotte Deogan, Jonathan E. Suk, Piotr Kramarz, Favelle Lamb, Pasi Penttinen

**Affiliations:** School of Medicine, University of Crete, Heraklion, Crete, Greece; Department of Oral Health Policy and Epidemiology, Harvard School of Dental Medicine, Harvard University, Boston, MA, USA; Division of Infection, Immunity and Respiratory Medicine, School of Biological Sciences, The University of Manchester, Manchester, UK; North West Lung Centre, Wythenshawe Hospital, Manchester University NHS Foundation Trust, Manchester Academic Health Science Centre, Manchester, UK; Department of Clinical Therapeutics, National and Kapodistrian University of Athens, Athens, Greece; Centre for Evidence Based Healthcare, Division of Epidemiology and Public Health, School of Medicine, University of Nottingham, UK; Heidelberg Institute of Global Health, University of Heidelberg, Germany; Catalan Institute of Oncology, Barcelona, Spain; Institut d’Investigació Biomèdica de Bellvithe (IDIBELL), Barcelona, Spain; European Centre for Disease Prevention and Control (ECDC), Sweden

## Abstract

**Background:** As mortality from COVID-19 is strongly age-dependent, we aimed to identify population subgroups at an elevated risk for adverse outcomes from COVID-19 using age/gender-adjusted data from European cohort studies with the aim to identify populations that could potentially benefit from booster vaccinations.

**Methods:** We performed a systematic literature review and meta-analysis to investigate the role of underlying medical conditions as prognostic factors for adverse outcomes due to SARS-CoV-2, including death, hospitalisation, Intensive Care Unit (ICU) admission, and mechanical ventilation within three separate settings (community, hospital and ICU). Cohort studies that reported at least age and gender-adjusted data from Europe were identified through a search of peer-reviewed articles published until 11^th^ June 2021 in Ovid Medline and Embase. Results are presented as Odds Ratios (ORs) with 95% confidence intervals (95%C.I.) and absolute risk differences (RD) in deaths per 1,000 COVID-19 patients.

**Findings:** We included 88 cohort studies with age/gender adjusted data from 6,653,207 SARS-CoV-2 patients from Europe. Hospital-based mortality was associated with high and moderate certainty evidence for solid organ tumours, diabetes mellitus, renal disease, arrhythmia, ischemic heart disease, liver disease, and obesity, while a higher risk, albeit with low certainty, was noted for chronic obstructive pulmonary disease and heart failure. Community-based mortality was associated with a history of heart failure, stroke, diabetes, and end-stage renal disease. Evidence of high/moderate certainty revealed a strong association between hospitalisation for COVID-19 and solid organ transplant recipients, sleep apnoea, diabetes, stroke, and liver disease.

**Interpretation:** The results confirmed the strong association between specific prognostic factors and mortality and hospital admission. Prioritisation of booster vaccinations and the implementation of non-pharmaceutical protective measures for these populations may contribute to a reduction in COVID-19 mortality, ICU and hospital admissions.

**Funding:** European Centre for Disease Prevention and Control (ECDC) under specific contract No. 10 ECD.11843 within Framework contract ECDC/2019/001 Lot 1B.

## INTRODUCTION

The COVID-19 pandemic has led to detrimental consequences for society, the global economy, and public health. Health care systems worldwide continue to face high pressure, particularly at peaks of transmission waves. After a period of decrease during the European summer months of 2021, an increase in the number of cases, hospital and Intensive Care Unit (ICU) admissions have been noted from October 2021 in most of the European Union and European Economic Area (EU/EEA) countries (1). According to the European Centre for Disease Prevention and Control (ECDC)’s most recent risk assessment, this rise was mainly attributed to the newly emerging variants of SARS-CoV-2, such as the Delta variant (B.1.617.2) and the Omicron variant (B.1.1.529), the loosening of non-pharmaceutical interventions (NPIs) across Europe and inadequate vaccination coverage (2).

In an effort to stratify patients with SARS-CoV-2 for better management of economic and human resources, particular attention has been paid to the determination of risk factors that predispose to adverse outcomes related to COVID-19, including hospitalisation and ICU admission or the need for ventilatory support, or death. Consistent evidence demonstrates a high risk of severe COVID-19 manifestations in older individuals, especially in combination with pre-existing medical conditions (3-5). Population-level data have indicated that COVID-19 older patients with comorbidities such as chronic cardiac, non-asthmatic chronic pulmonary, chronic kidney and liver diseases, and obesity have higher mortality in hospitals (6-8). Although the European Surveillance System (TESSy) has noted through the use of population data, that COVID-19 patients with cardiac disorders (25·7%), diabetes (15·5%), and cancers (9·9%) have the highest case-fatality rates in the European population (9), and while previous systematic reviews have separately assessed several clinical indicators or comorbidities, they lack age/gender-adjusted analyses and stratification by patient setting (4).

As mortality from COVID-19 is strongly age-dependent, a meta-analysis of pooled age-adjusted estimates from available cohort studies is needed to determine which comorbidities should classify patients into high-risk groups for adverse COVID-19 outcomes. Such evidence would be of interest to clinicians to better manage patient flow and to policymakers when planning forthcoming public health measures, such as booster vaccination strategies and personal NPIs in coming phases of the pandemic.

## METHODS

The systematic review as conducted adhered to the PRISMA (Preferred Reporting Items for Systematic Reviews and Meta-Analysis) (10) and MOOSE (Meta-analyses Of Observational Studies in Epidemiology) guidelines (11). The protocol of this systematic review was pre-reviewed by the European Centre for Disease Prevention and Control (ECDC).The protocol was not pre-registered in any database for systematic reviews.

Cohort studies were considered eligible provided (i) they evaluated patients with clinically diagnosed or laboratory-confirmed COVID-19 in one of the following settings, each reflecting different disease stages or severity: community, hospitalised patients, intensive care unit (ICU); (ii) they were conducted in Europe, including EU/EEA countries, the United Kingdom (UK) and Switzerland; (iii) they assessed the association between underlying clinical conditions and the primary adverse outcomes of COVID-19; iv) Any underlying clinical condition described in original studies was considered acceptable for inclusion in the meta-analysis, however the statistical analysis presented had to be at least age and gender-adjusted. The primary outcomes of our meta-analyses were mortality, hospital admission and ICU admission. For the secondary outcomes, we also assessed the use of mechanical ventilatory support and a composite outcome comprising admission to the ICU and/or death and/or hospice care (e.g., “Death or ICU admission”). The latter secondary outcome was considered crucial to account for the bias introduced by excluding patients from higher levels of care due to their poor baseline status or severe comorbidities.

Relevant peer-reviewed studies published in English were identified within Medline (OVID) and EMBASE (OVID) until the 11^th^ of June 2021. Subject headings relating to COVID-19 and epidemiological study design terms were used to develop a comprehensive search strategy presented in **Appendix 1**. Reference lists of all included studies and identified reviews were also screened to identify additional relevant studies. Full texts of potentially eligible studies were evaluated independently by two reviewers. Disagreements or uncertainties in screening stages were resolved through discussion and consensus.

An ad hoc designed structured form was used for extracting relevant data from each eligible study, including details on the study design, baseline characteristics of the participants and relevant outcome data. For accuracy, each study’s data were extracted by one reviewer, with each study cross-checked by a second reviewer. Adequate information was extracted to allow us to identify overlapping populations. In the case of overlapping populations, we prioritised including data from the study with the larger population and a more rigorously described methodology.

The methodological quality of each included study was evaluated independently by two reviewers using the Joanna Briggs Institute (JBI) standardised critical appraisal tool for the appropriate design (8). Disagreements were resolved with discussion and, when necessary, adjudication by a third reviewer.

### The certainty of the evidence

The Grading of Recommendations Assessment, Development and Evaluation (GRADE) methodology was used for evaluating the certainty in the body of evidence for each meta-analysis (12). In line with GRADE recommendations for the assessment of evidence about prognostic factors, we initially ascribed high certainty for all our findings which were subsequently rated down in cases of study limitations of the included studies, inconsistency, indirectness or imprecision of the results, or evidence of publication bias. Moreover, certainty was rated up in cases of large observed effects. Results that were of high or moderate certainty are primarily reported in the text of the current article.

### Statistical Analysis

In anticipation of significant clinical and methodological heterogeneity in the meta-analyses, we fitted logistic regression models with random effects. All outcome data were adjusted at least for gender and age. All variables analysed were dichotomous and analysed as Odds Ratios (OR) with corresponding 95% confidence intervals (95% C.I.). Heterogeneity was quantified using the I^2^ statistics. We considered values above 75% to represent considerable heterogeneity. To facilitate interpretability of the results and in line with recommendations by GRADE (12), we also present the absolute Risk Differences (RD) per 1,000 COVID-19 patients with corresponding 95% C.I. The median prevalence of the evaluated risk factors and the median incidence rate of each adverse outcome in each recruitment setting were used for calculating the absolute RDs.

## RESULTS

We included 88 studies, involving 6,653,207 cases with COVID-19 in the review (**PRISMA flowchart: Figure 1**). Recruitment dates ranged from January 1st, 2020, to April 30^th^, 2021, but were primarily based in 2020. There were 69 cohort studies that assessed 6,029,126 patients in the hospital setting (13-81), 13 cohort studies involving 621,792 patients in the community setting -of which ten were based solely in the community (82-91) while three combined community/hospital recruitment (92-94), and six cohort studies involving 2,289 patients were based in the ICU setting (95-100). Details on the characteristics of the included cohort studies are presented in **Appendix 2**, while the assessment of the risk of bias according to the JBI critical appraisal tool is provided in **Appendix 3**.

**Figure 1.**
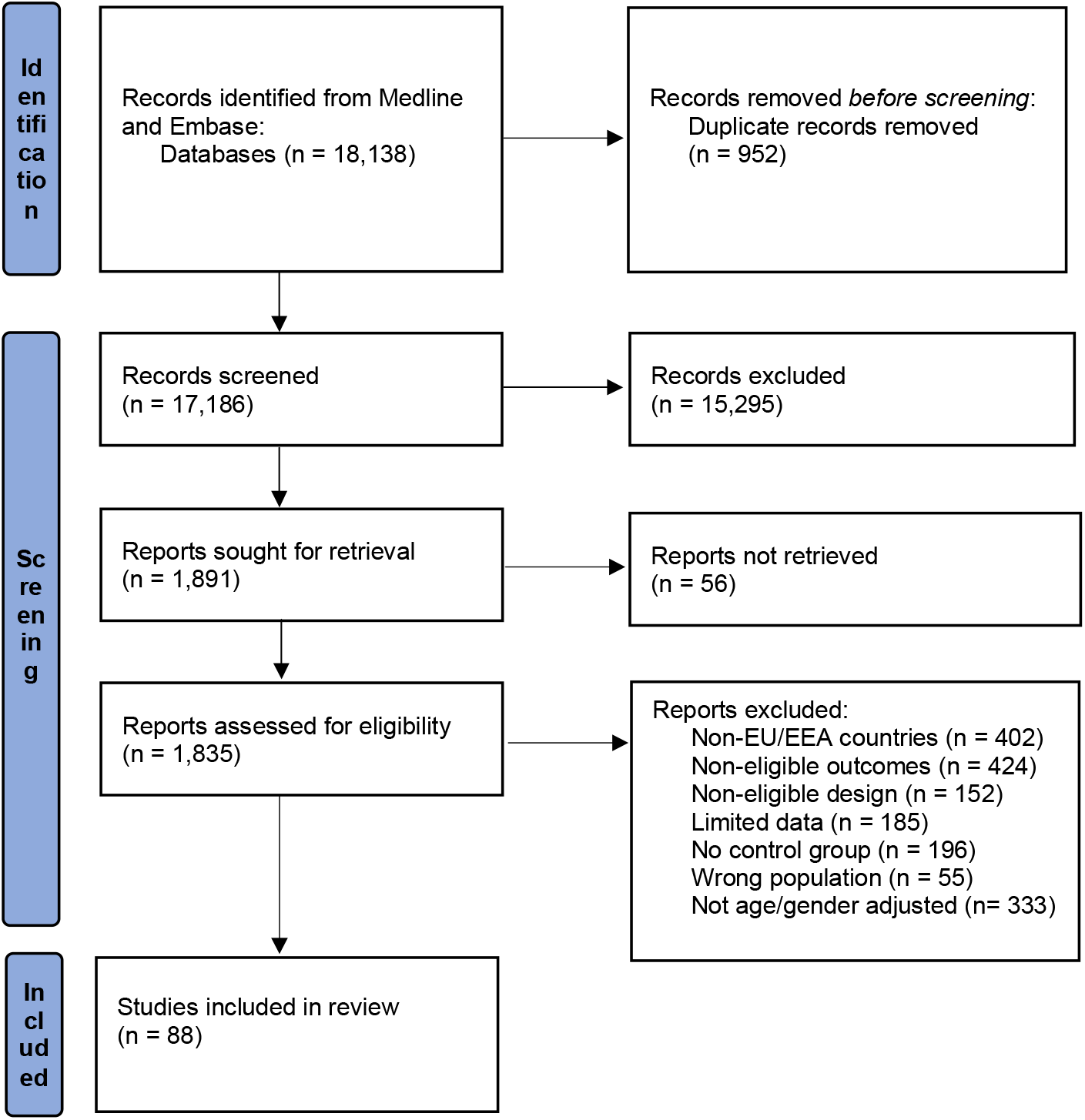
PRISMA Flowchart of the included studies.

As per our inclusion criteria, all cohort studies were based in Europe and included 23 studies from Spain (14, 16, 17, 22, 28, 29, 32, 33, 35, 50, 54, 57-59, 64, 67, 70, 80, 89, 92, 96, 97, 101), 22 studies from Italy(13, 19, 21, 23, 25, 30, 38, 39, 45, 47, 48, 55, 56, 61, 62, 68, 71, 77, 79, 90, 91, 93), 15 studies from France (20, 24, 27, 31, 40, 41, 43, 53, 60, 63, 65, 74, 76, 86, 99), ten studies from the UK (26, 34, 44, 49, 52, 66, 72, 75, 87, 98), four included patients from more than one country (69, 73, 88, 94), three from Germany (46, 51, 78), two studies from Sweden(36, 100), two from Denmark (42, 83) and one study from Belgium (81), Finland (5), the Netherlands (18), Norway (85) and Poland (37). The diagnosis of COVID-19 was made mainly with a PCR test, except for in five studies in which the diagnosis was made with the ICD Classification (20, 24, 27, 42, 102).

### Underlying medical conditions as prognostic factors of mortality due to COVID-19

While all meta-analyses findings are presented with both adjusted OR (aOR) and absolute risk differences (RD) in **Figures 2a-b**, below, we primarily present key results supported by evidence of moderate or high certainty as assessed using the GRADE approach.

**Figure 2:**
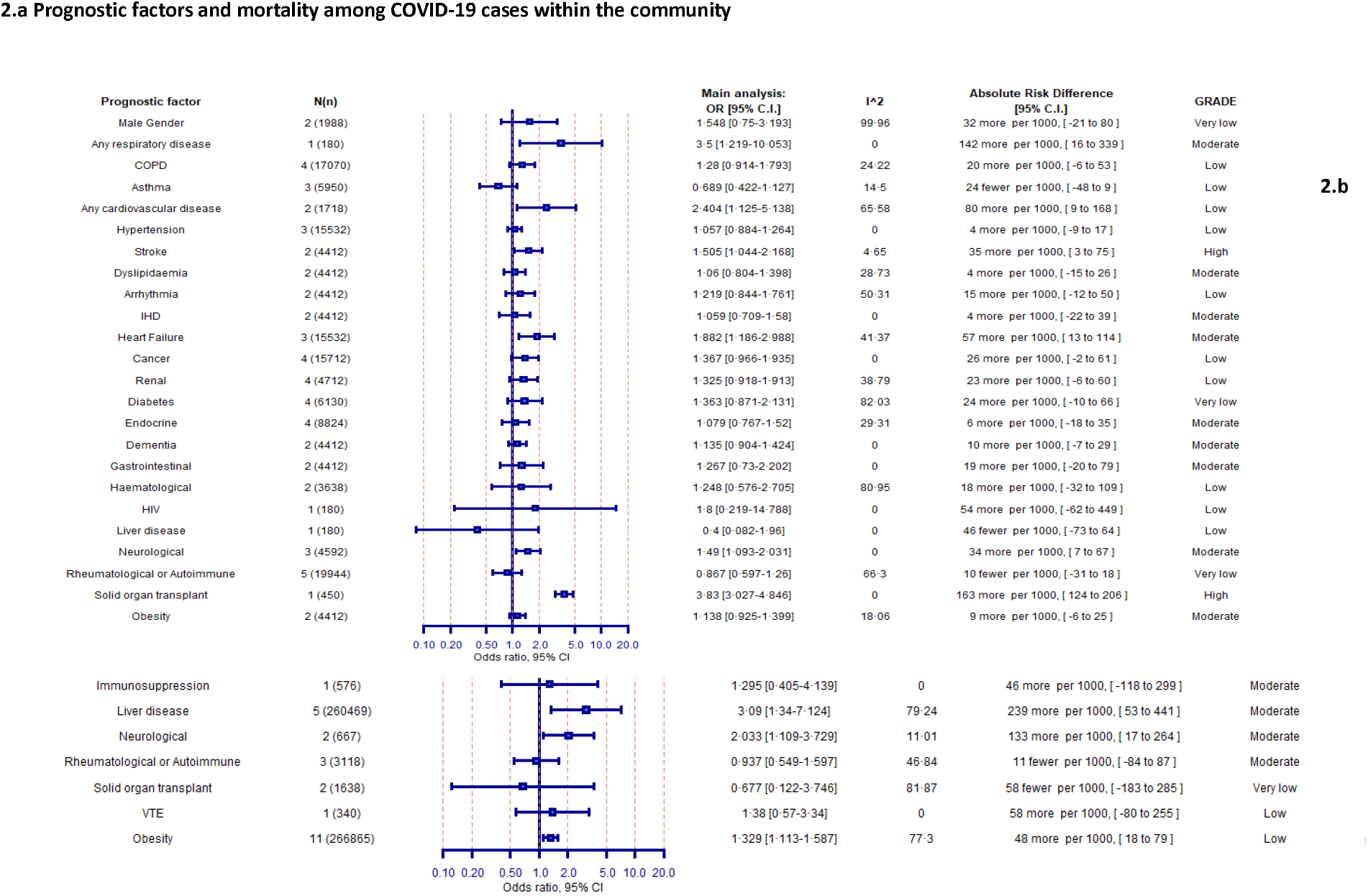
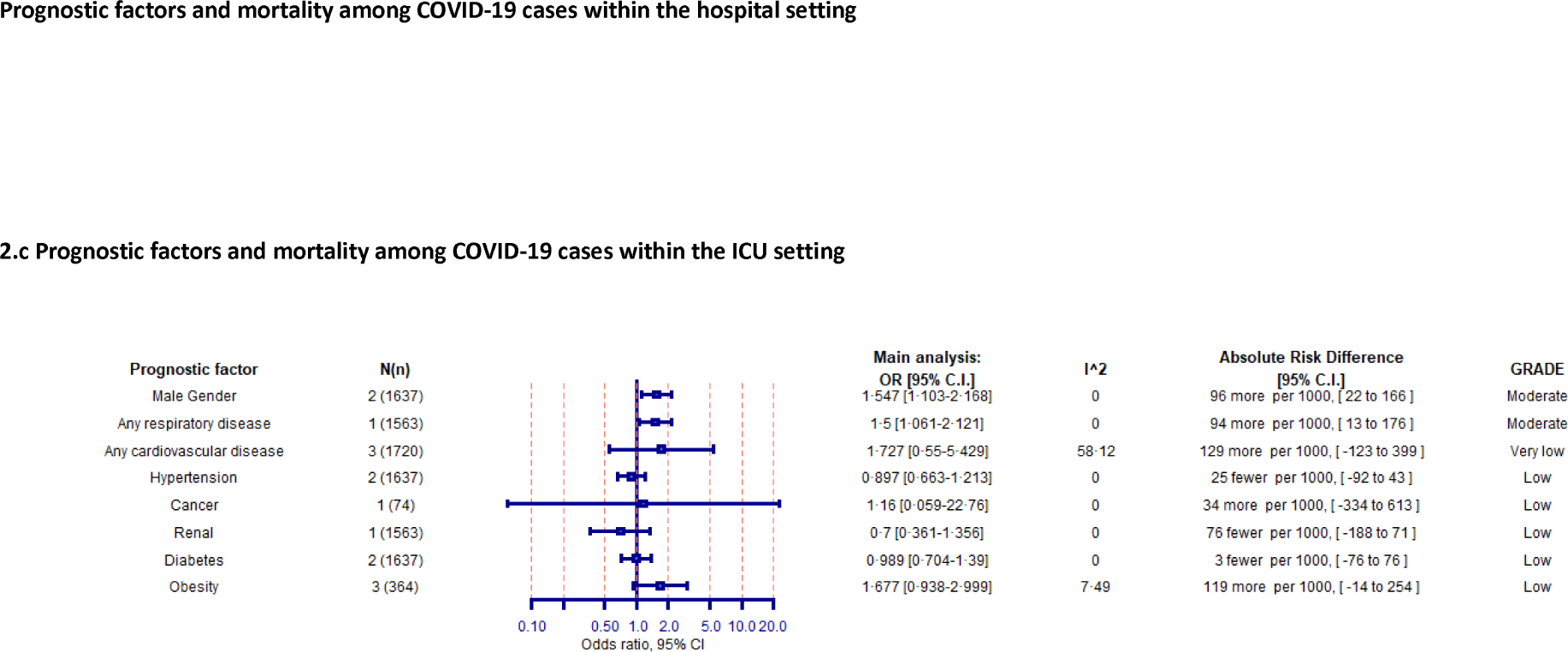
Association between prognostic factors and mortality among COVID-19 cases within the community (2a), hospital (2c) and ICU settings (2d). Odds ratios (95% confidence intervals), I^2 test for heterogeneity, absolute risk differences (95% confidence intervals) and the GRADE assessment are presented

The presence of any cardiovascular disease posed an increased risk for mortality in the hospital setting (51 [17-85] more deaths per 1,000 cases; high certainty). Additionally, stroke (35 [3-75] more deaths per 1,000 cases; high certainty) and heart failure (57 [13-144] more deaths per 1,000 cases; moderate certainty) were associated with increased mortality risk in the community setting, while ischaemic heart disease (IHD) was estimated to increase the risk of death in hospitalised patients with COVID-19 by 187 [11-385] more deaths per 1,000 cases (moderate certainty). Diabetes mellitus was associated with increased mortality in the hospital setting as 57 [31-84] (high certainty) additional deaths were observed for every 1,000 patients recruited with COVID-19 and diabetes mellitus. Arrythmia was associated with increased mortality in the hospital setting (174 [28-338]; moderate certainty). A history of solid organ transplant receipt was associated with increased mortality in the community setting (163 [124-206] more deaths per 1,000 cases; high certainty]). Patients with neurological disease were also at an elevated risk of death within both the hospital (133 [17-264] more deaths per 1,000 cases; moderate certainty) and the community setting (34 [7-67] more deaths per 1,000 cases; moderate certainty). Similarly, COVID-19 patients with any respiratory disease were found to have increased mortality risk in the community (142 [16-339] more deaths per 1,000 cases; moderate certainty) and in ICU (94 [13-176] more deaths per 1,000 cases; moderate certainty) setting.

Other factors associated with an increased mortality in the hospital setting were renal disease (91 [25-163]; moderate certainty); dementia (99 [4-209]; moderate certainty); cancer (121 [78-166]; high certainty); liver disease (239 [53-441]; moderate certainty); and male gender (68 [50-86]; high certainty). Male gender was found to be associated with increased death also in the ICU setting (96 [22-166]; moderate certainty). Data regarding the presence of other comorbid diseases with COVID-19 mortality were less consistent or of lower certainty.

### Hospital and ICU admission

**Figure 3** presents the Forest plot that summarises the available evidence presented as aOR and absolute RDs regarding the association between COVID-19 and hospital admission within the community setting. Increased hospitalisation per 1,000 COVID-19 patients was noted for the history of solid organ transplant [320; 276-360] and sleep apnoea [262; 5-457], supported by evidence of high certainty. Moreover, other prognostic factors were associated with hospital admission with evidence of moderate certainty and included any cardiovascular disease [156; 67-242]; heart failure [291; 190-379]; any respiratory disease [278; 157-380]; diabetes [129; 73-185] and male gender [137; 81-192]. **Figure 4** presents the available evidence of the association between prognostic factors and ICU admission within the hospital setting due to COVID-19. Supported by evidence of moderate certainty, the presence of asthma and neurological diseases was significantly associated with ICU admission, other factors were supported by evidence of lower certainty or indicated non statistically significant results.

**Figure 3.**
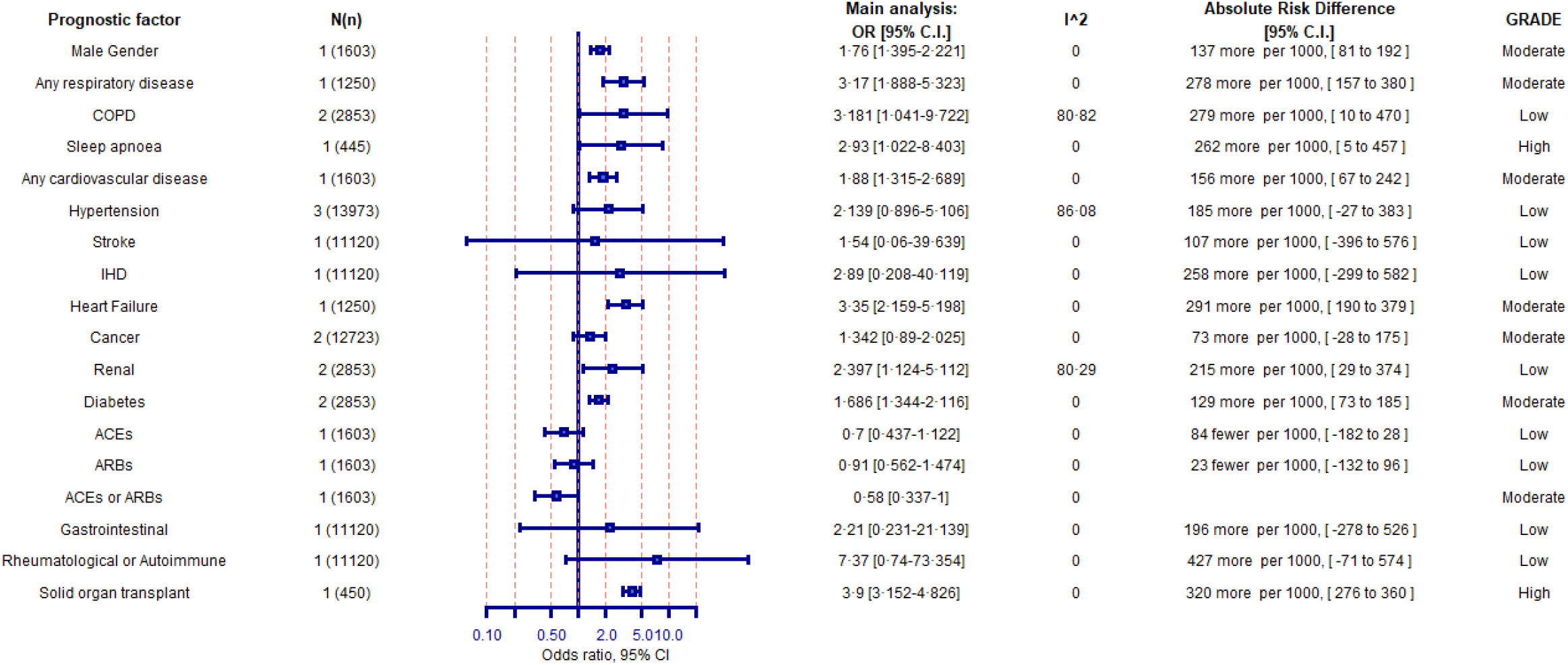
Association between prognostic factors and hospital admission among COVID-19 cases within the community setting. Odds ratios (95% confidence intervals), I^2 test for heterogeneity, absolute risk differences (95% confidence intervals) and the GRADE assessment are presented for all studies

**Figure 4.**
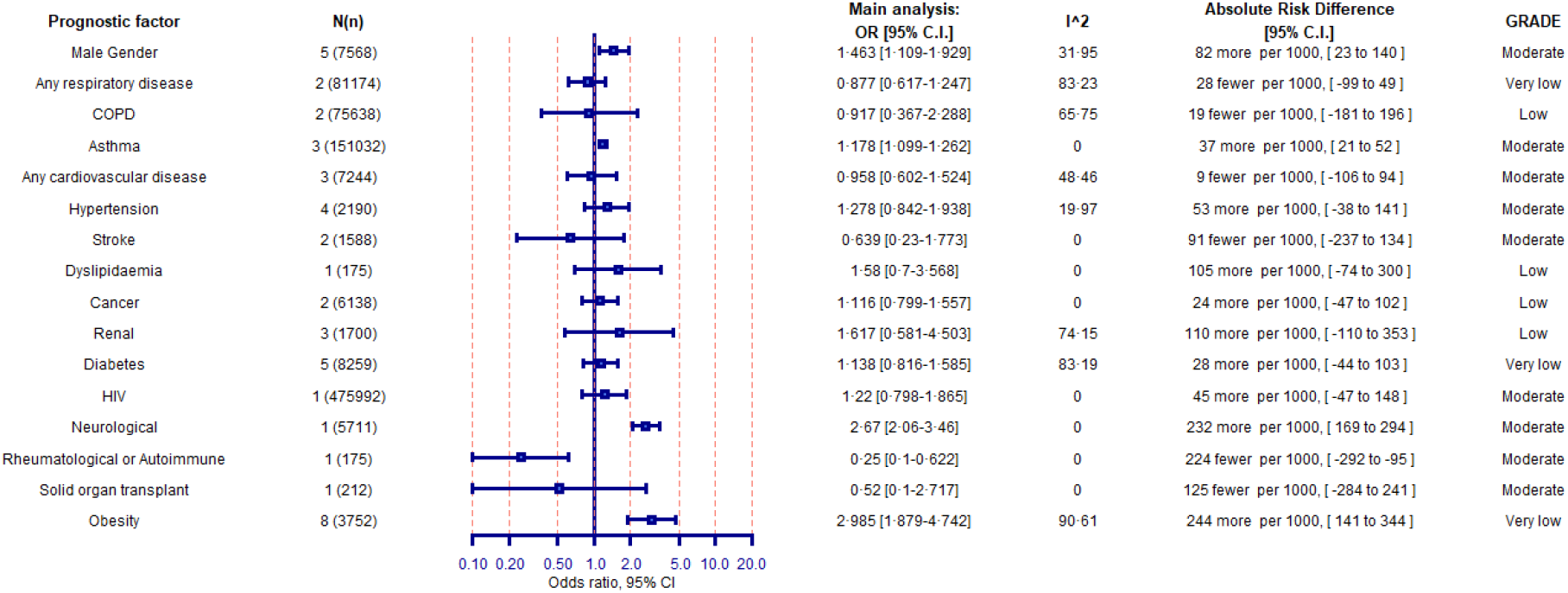
Association between prognostic factors and ICU admission among COVID-19 cases within the hospital setting. Odds ratios (95% confidence intervals), I^2 test for heterogeneity, absolute risk differences (95% confidence intervals) and the GRADE assessment are presented for all studies

### Composite outcome of death or ICU admission

As a supplementary analysis, we assessed the prognostic factors related to the composite adverse outcome of combined death and/or ICU admission in **Appendix 4** for the community (**Appendix 4a**) and the hospital setting (**Appendix 4b**). Among hospitalised patients, the presence of COPD [252; 115-383]; dyslipidaemia [95;4-190]; hypertension [91;27-153]; diabetes [71;47-95] and any cardiovascular disease [76; 1-153] was associated with this composite outcome supported by evidence of high certainty. Moreover, obesity [143;94-192]; heart failure [141;39-246] and male gender [144; 98-188] were significantly associated with this composite outcome in the hospital setting, supported by evidence of moderate certainty. Diabetes [33; 4-66] and male gender [33; 12-54] were also estimated to increase the risk for ICU admission or death in the community setting with evidence of moderate certainty.

### Mechanical ventilation

***Appendix 5*** presents the assessment of the need for mechanical ventilation, as a secondary outcome, for both the hospital setting (**Appendix 5a**) and the ICU setting (**Appendix 5b**). Obesity was associated with an increased need for mechanical ventilation in both the hospital [169 (162-176); high certainty] and ICU setting [128 (67-176); low certainty]. Moreover, data of high certainty indicated an increased risk for mechanical ventilation in hospitalised COVID-19 patients with asthma [18; 2-35], while male gender was associated with higher odds of mechanical ventilation need in both hospital [108 (48-164); high certainty] and ICU setting [172 (3-321); moderate certainty].

## DISCUSSION

Based on quantitative age and gender-adjusted data extracted from 88 cohort studies conducted in Europe, reporting on 6,653,207 patients with COVID-19, this systematic review and meta-analyses strengthen the understanding of prognostic factors for adverse outcomes of infection that would set the base for the identification of high-risk populations in order to support public health decision making regarding booster vaccinations and personal non-pharmaceutical interventions as societies re-open across Europe.

Extensive research has confirmed the strong association of age and gender with mortality and hospital admissions (103-107) hence it is important to take these factors into account in meta-analyses, particularly given the strong association of COVID-19 outcomes with age, which likely leads to an overestimation of the risk posed by ageing-associated diseases, such as congestive heart disease, COPD, or dementia. Hence, in contrast to previous meta-analyses, our approach evaluated the weight of evidence for factors that were at least age/gender adjusted to overcome the issue. Notably, male gender was identified as a strong predisposing factor across multiple analyses even after adjusting for patient age.

### Hospital and ICU Mortality

In our age/gender-adjusted analyses, increased hospital mortality was noted among patients with COPD, arrhythmia, IHD, heart failure, cancer, renal disease, liver disease, obesity and diabetes. Previous systematic reviews had identified COPD, cardiovascular disease, hypertension, diabetes, chronic renal disease and chronic liver disease as factors associated with mortality in the hospital setting, however within these reviews, substantial heterogeneity was noted, and their results were unadjusted (107-110). With regards to ICU mortality, our meta-analysis of adjusted data for ICU mortality due to COVID-19 indicated that male gender and the presence of respiratory disease was associated with mortality. However, unadjusted meta-analyses showed associations with several comorbidities, such as COPD and renal disease – results also identified in a recent meta-analysis of unadjusted data on ICU patients which identified hypertension, COPD and cardiovascular disease as prognostic factors of mortality (111). Whilst obesity has been identified in previous meta-analyses to be strongly associated with mortality (112), within our meta-analyses, the data available for obesity were heterogeneous due to different classifications used within each study. Obesity was associated with overall hospital mortality, supported however only by very low quality of evidence.

### Hospital and ICU admission

Our meta-analysis focusing only on European data and evaluating ICU admission indicated that in age/gender-adjusted analyses, asthma, neurological diseases and autoimmune diseases were associated with an increased likelihood of the need for ICU admission. Furthermore, the need for mechanical ventilation was higher among patients with asthma, liver disease and obesity. These results are different from other ICU cohort data, which may be attributable to our analyses being age/gender adjusted (113), although previous research has also indicated that ICU outcomes may be better predicted by frailty than either age or comorbidity (114). Notably, our analyses noted that several comorbidities associated with an increased risk of death or hospital admission were not typically associated with an increased incidence of ICU admission or mechanical ventilation. This is not unexpected since patients with pre-existing severe comorbidities may not be offered ICU admission or mechanical ventilation as therapeutic options during times of unprecedently high pressure on healthcare systems, leading to strict triage criteria (115, 116).

COVID-19, during the first wave of the pandemic, was managed heterogeneously worldwide, especially in resource-constrained countries, where the health systems have often had less capacity and resources to face the unfolding pandemic (117). However, even in the selected high-income countries, ICUs were under unprecedented pressure, and strict criteria for selecting patients were applied based on the patients’ short-term prognosis (118).

The above results indicate populations of high risk for adverse outcomes and could possibly benefit from potential booster vaccinations, as a preventive measure to reduce their elevated risk for adverse COVID-19 outcomes. However, our analyses can also assist informing planning of patient flows in the health care settings. Notably, outcomes and prognostic factors in different settings were often different, a factor which may be attributable to several factors. It is possible that the fact that the majority of those contracting SARS-CoV-2 experience a mild or asymptomatic disease, and only a minority progresses to the development of pneumonitis, which may warrant hospital admission (119). Another potential explanation may be the variability of the impact of different comorbidities on disease progression or clinician bias to the presence of specific comorbidities among patients triaged and on the threshold of hospital admission (e.g., a patient with COVID and concomitant COPD is more likely to be admitted than a patient without COPD), indicating a type of selection bias by the clinicians during triage.

### Strengths and limitations

Several strengths of our meta-analysis increase our confidence in our findings. The inclusion of only age/gender adjusted cohort study data allowed us to control for the primary factor associated with adverse outcomes, which is the patient’s age, hence allowing us to assess the independent effect of each prognostic factor (106). Moreover, we performed separate analyses by clinical setting that allowed us to both quantify the incidence of adverse outcomes, as separate endpoints, which may assist in managing patient flows and triaging. As we applied the GRADE methodology for assessing the evidence about prognostic factors, we rigorously evaluated the certainty of the evidence behind each meta-analysis. Some limitations of our study should also be acknowledged as our results reflect the status quo in Europe during the first waves of the pandemic when the alpha and beta variants were most dominant and with the EU population largely unvaccinated, hence further research is needed to assess how subsequent vaccination would impact patient mortality. We restricted our inclusion criteria to Europe, to enhance the comparability of the results and utility for European clinicians and policymakers, as they have similar healthcare resources, acknowledging that our results may not be generalisable to other areas of the globe. However, similar results are noted in different geographical areas (107). Furthermore, there was evidence of heterogeneity as the definition of specific prognostic factors, such as hypertension, obesity, and dyslipidaemia, were ambiguous in some of the included studies and variability in the definitions may have also contributed to the observed variability.

Given that a direct comparison among different risk factors was not performed and similar effect sizes were found, no conclusions can be drawn on the comparative ranking or cumulative/synergistic effect of specific prognostic factors leading to COVID-related adverse outcomes. Nevertheless, conclusions on the individual impact of specific comorbidities can be drawn.

## CONCLUSION

The results of this systematic literature review and meta-analysis of European cohort age/gender adjusted data indicate that COPD, arrhythmia, IHD, heart failure, cancer, renal disease, liver disease, obesity and diabetes were associated with hospital mortality while male gender and respiratory diseases were associated with ICU mortality. COPD, dyslipidaemia, hypertension, diabetes, cardiovascular disease, obesity, heart failure and male gender were associated with the composite outcome of death and/or ICU admission. With regards to patient management, factors that were associated with hospital admission included solid organ transplant, sleep apnoea, cardiovascular disease, diabetes, heart failure, respiratory disease and male gender while obesity, asthma and male gender were associated with the need for mechanical ventilation. Taking the above into account, the prioritisation of preventive public health measures, such as potential booster vaccinations and use of personal NPIs, could be further refined by accounting for underlining pre-existing conditions as societies and health care systems across Europe continues to face pressure from COVID-19.

## Data Availability

All data produced in the present work are contained in the manuscript

## FUNDING - ACKNOWLEDGEMENTS

### Funding

This report was commissioned by the European Centre for Disease Prevention and Control (ECDC) to the PREP-EU Consortium, coordinated by Dr Constantine Vardavas (School of Medicine, University of Crete) under specific contract No. 10 ECD.11843 within Framework contract ECDC/2019/001 Lot 1B.

### Role of the Funding Agency

The funder contributed to the definition of the scope of the review and commented on the protocol and draft manuscript and was not involved in the conduct of the systematic review. ECDC provided intellectual input throughout the project and arranged for an internal peer review of the final manuscript prior to submission. The lead author has full access to the data and has the responsibility for submission.

### Contributors

CV, JL-B, RP and JES designed the study. KN, DC-P and AGM undertook the literature review and extracted the data. JL-B and RP developed the search code. AGM, JV, KS and KN analysed and interpreted the data. EF and GG participated in data evaluation and interpretation along with CV, JL-B, RP, JES, KN, KS, and AGM. CV wrote the first draft of the manuscript with input from all authors. JS, CD, PK, FV and PP along with all other authors reviewed and revised subsequent drafts.

## Acknowledgements

We would like to thank Katerina Papathanasaki (UoC), Chrysa Chatzopoulou (UoC) and Konstantinos Skouloudakis (UoC) for their assistance in data archiving. We acknowledge the bravery and dedication of healthcare professionals everywhere during these most challenging times.

## Data sharing statement

Data sharing is not applicable to this article as no new data were created or analysed in this study. All data were extracted from peer-reviewed studies, which were identified from the literature and have been appropriately referenced within the article.

## APPENDIXES

## Appendix 1: Search strategies for identifying studies

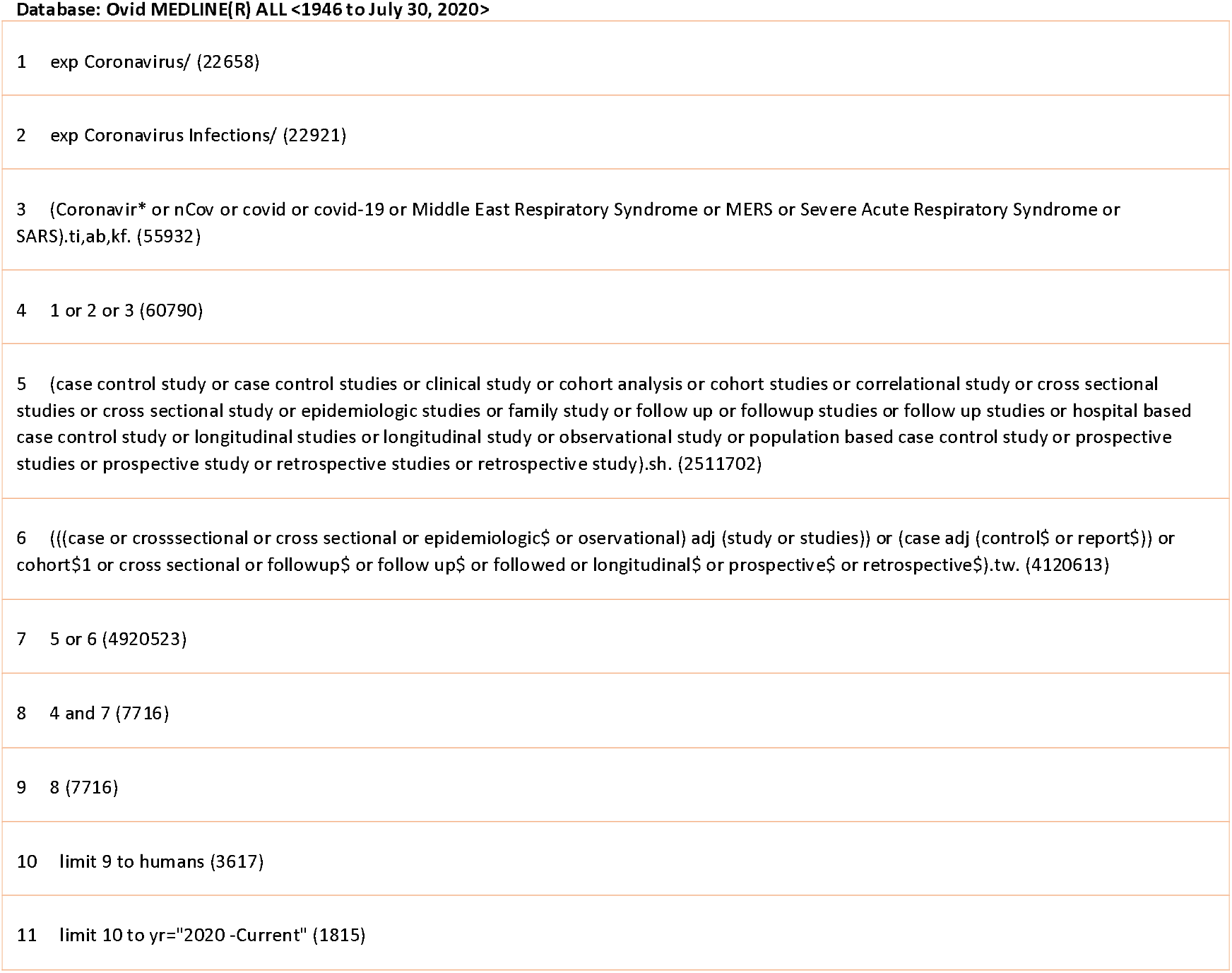
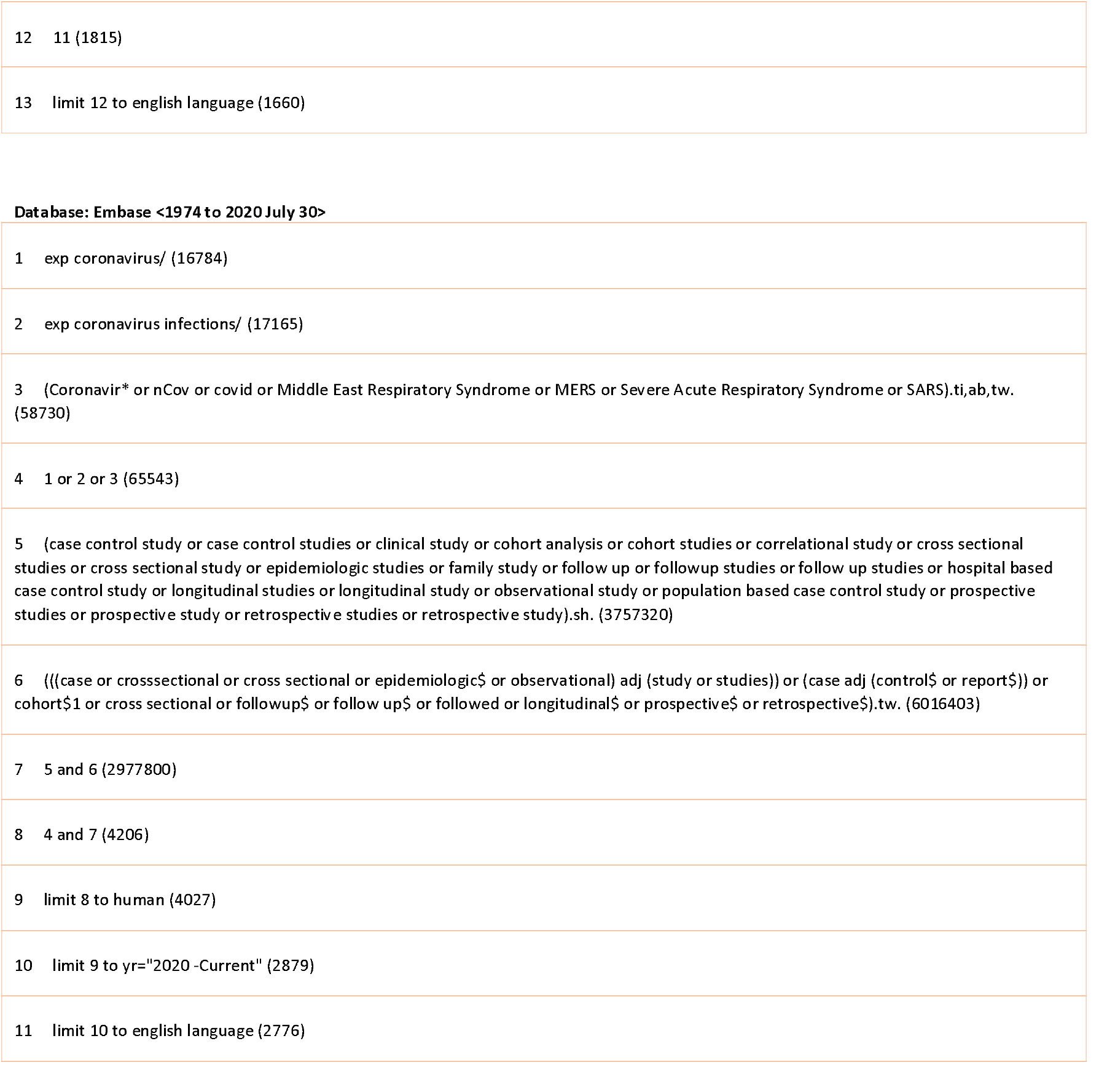

## Appendix 2: Characteristics of the included studies

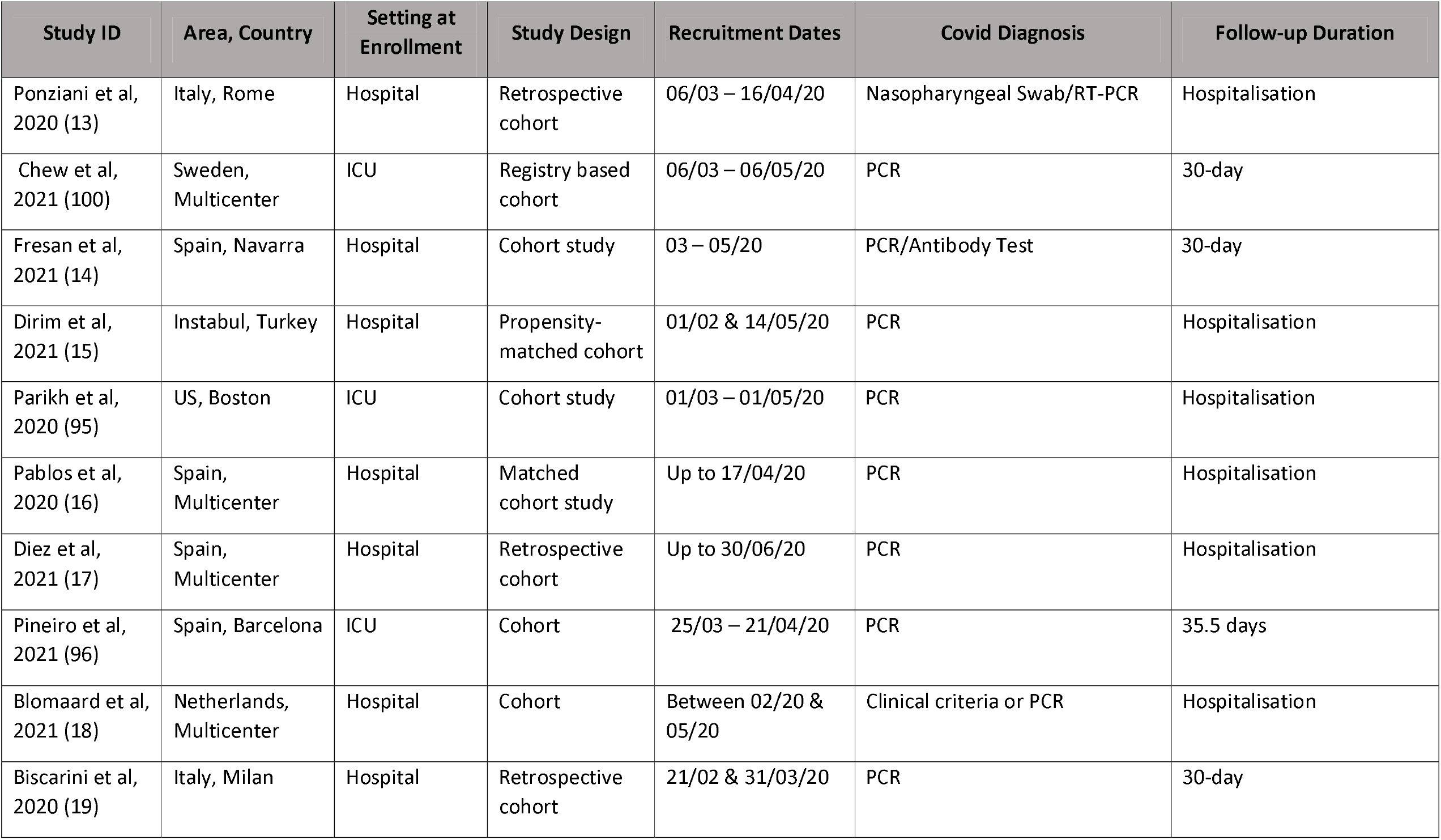
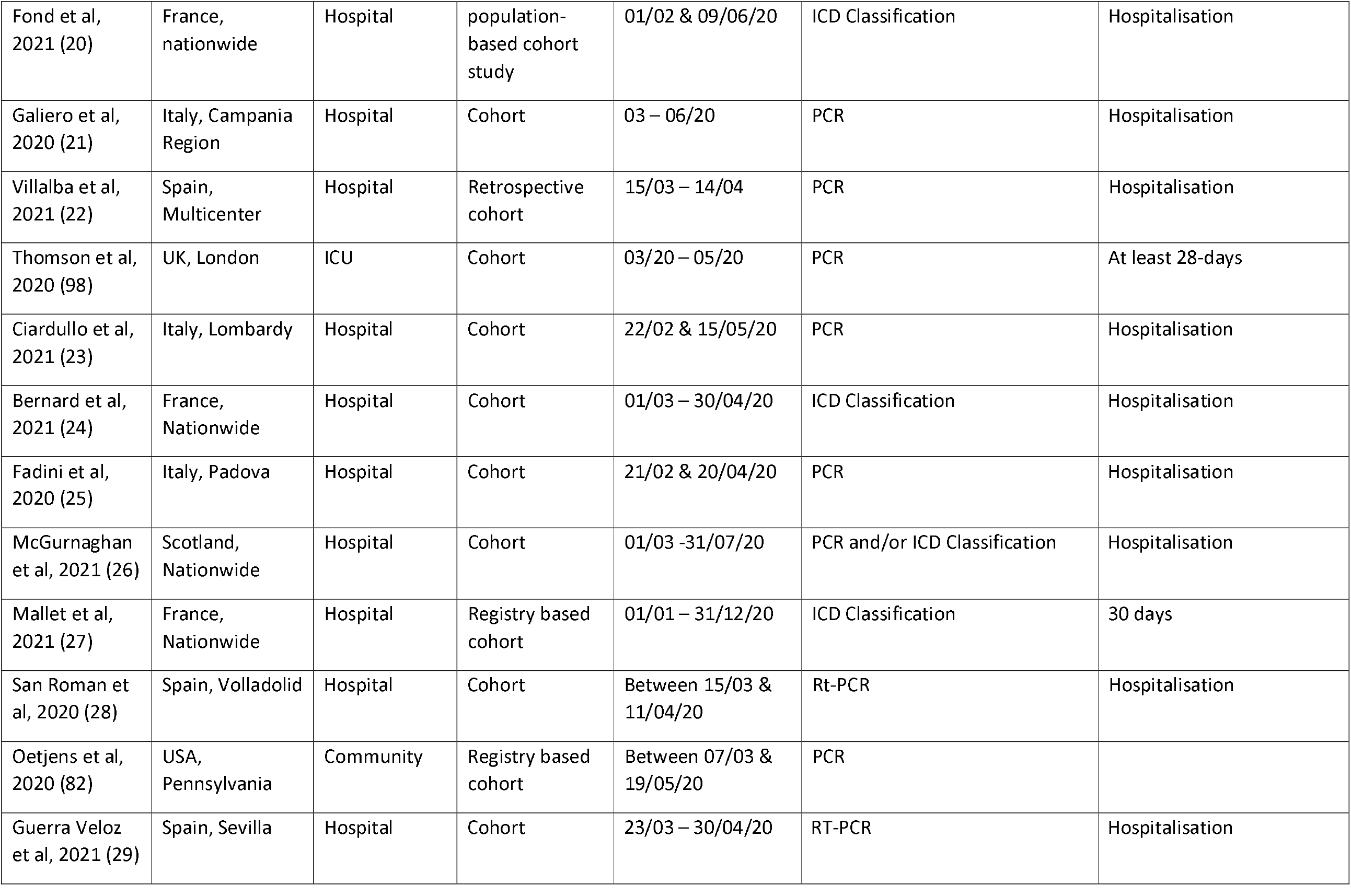
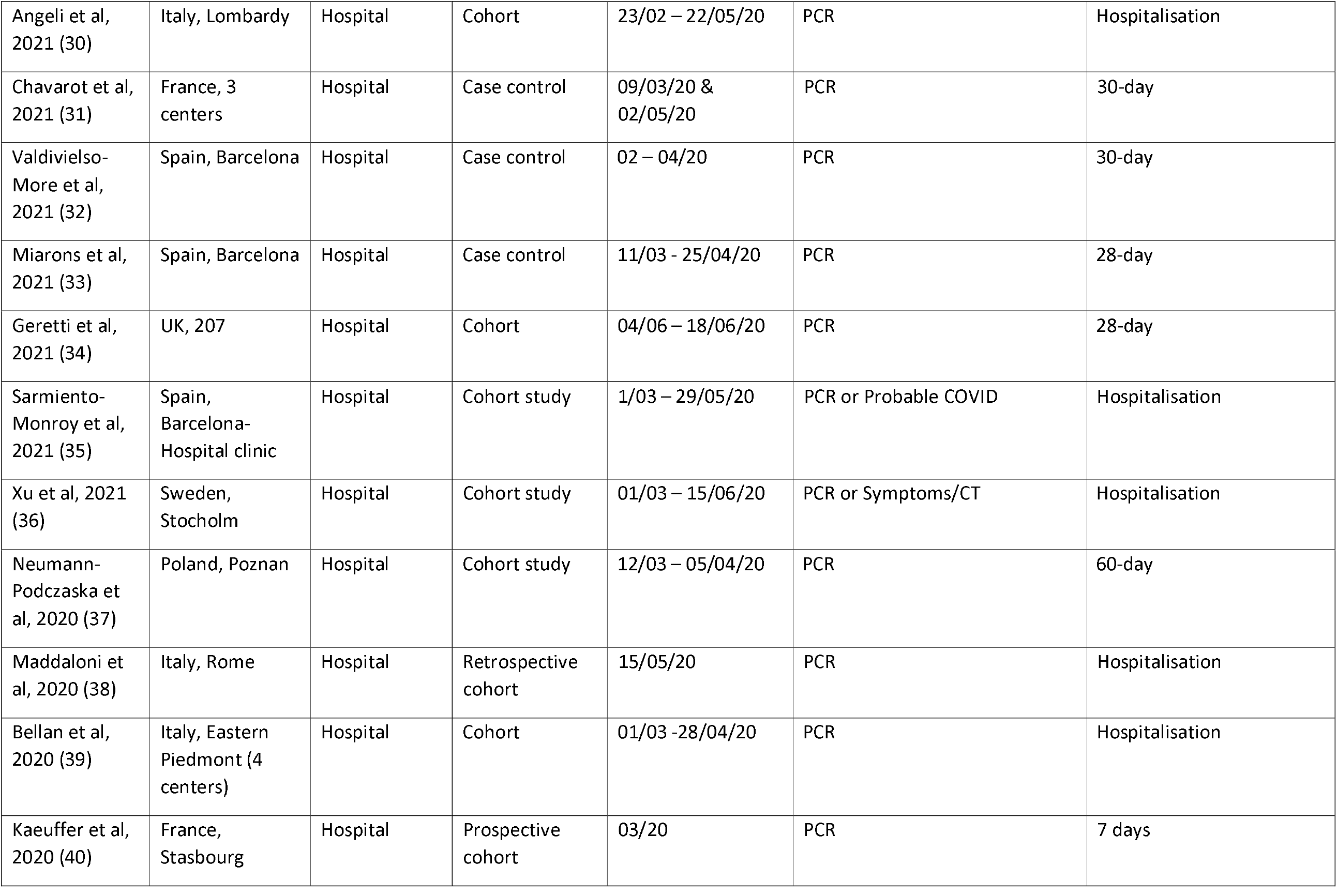
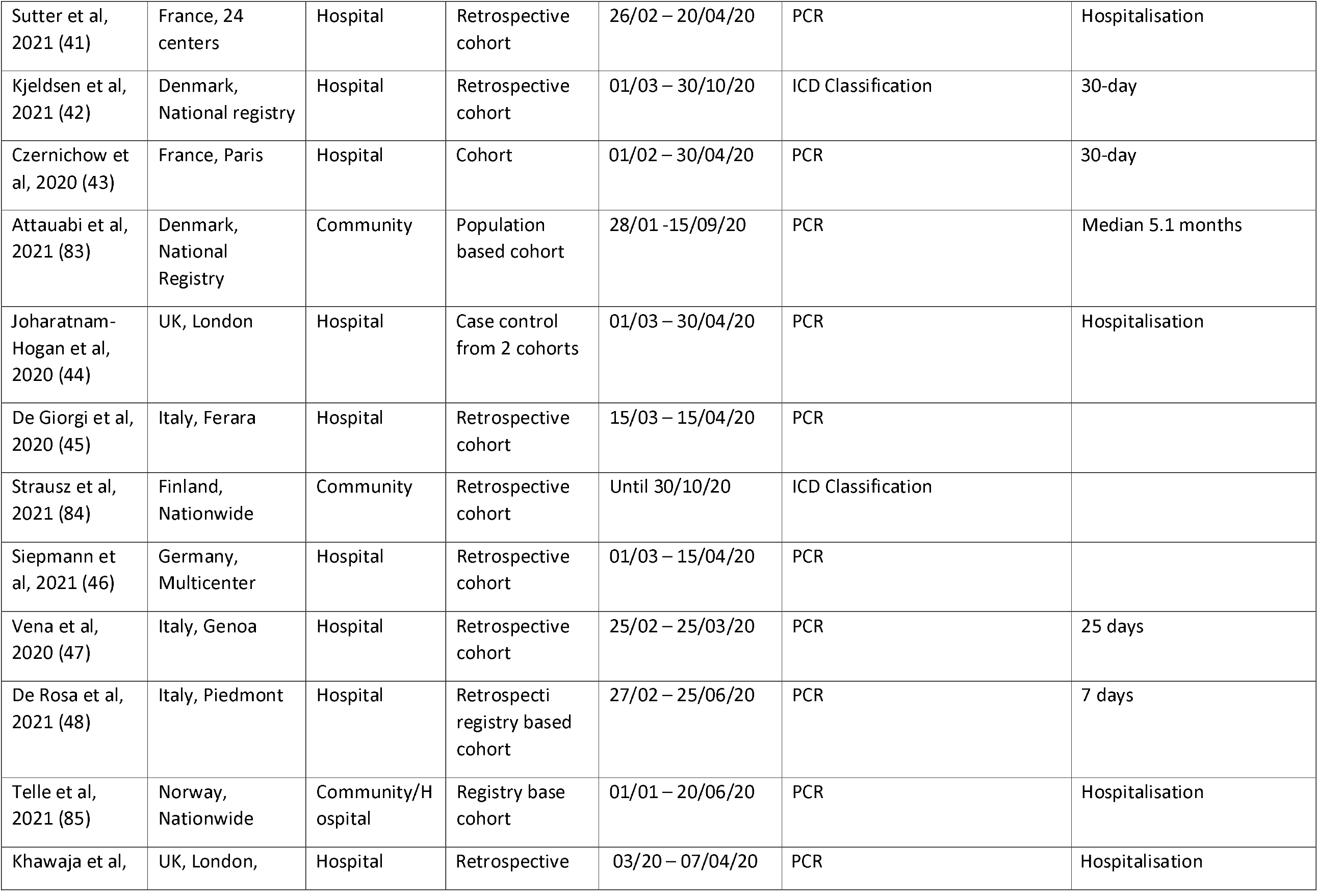
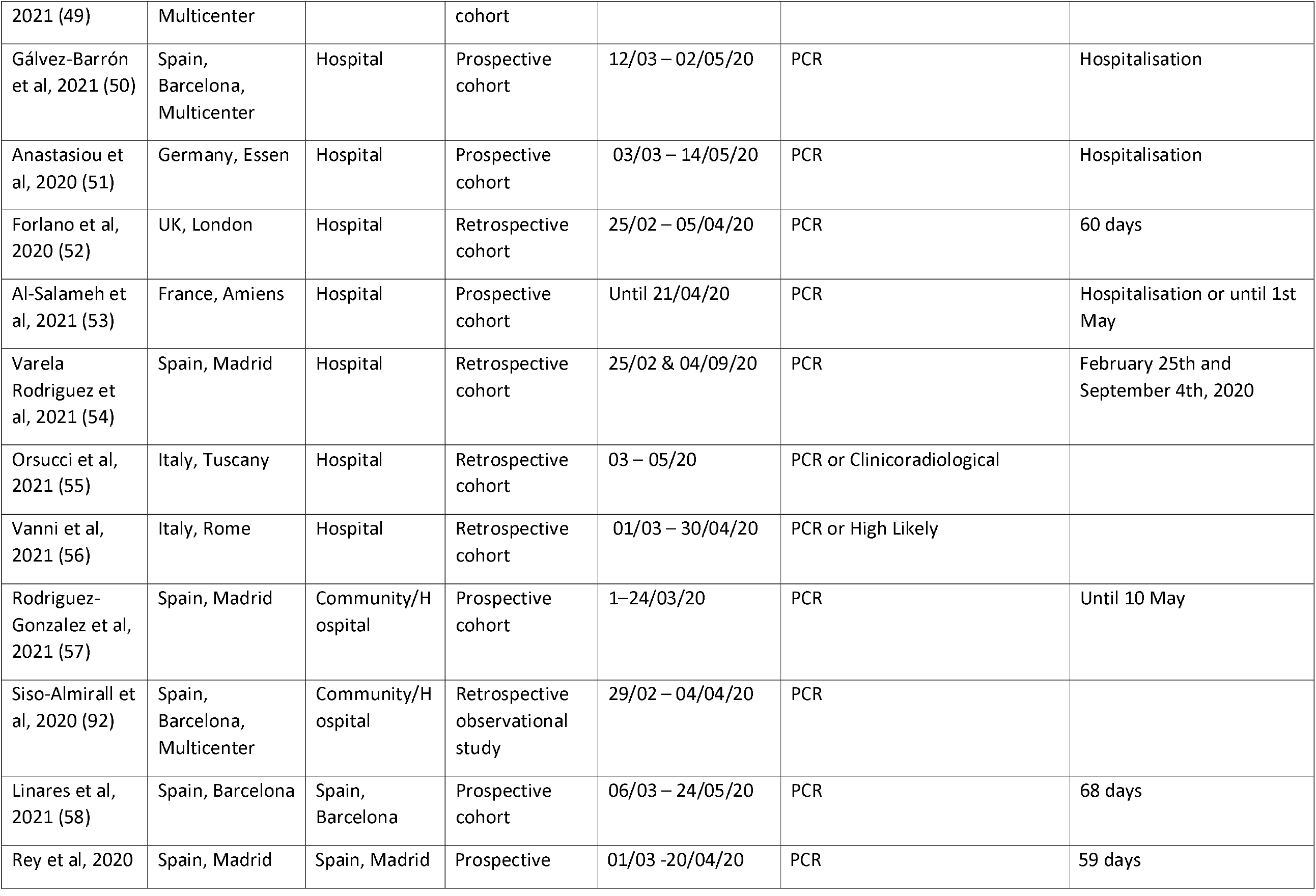
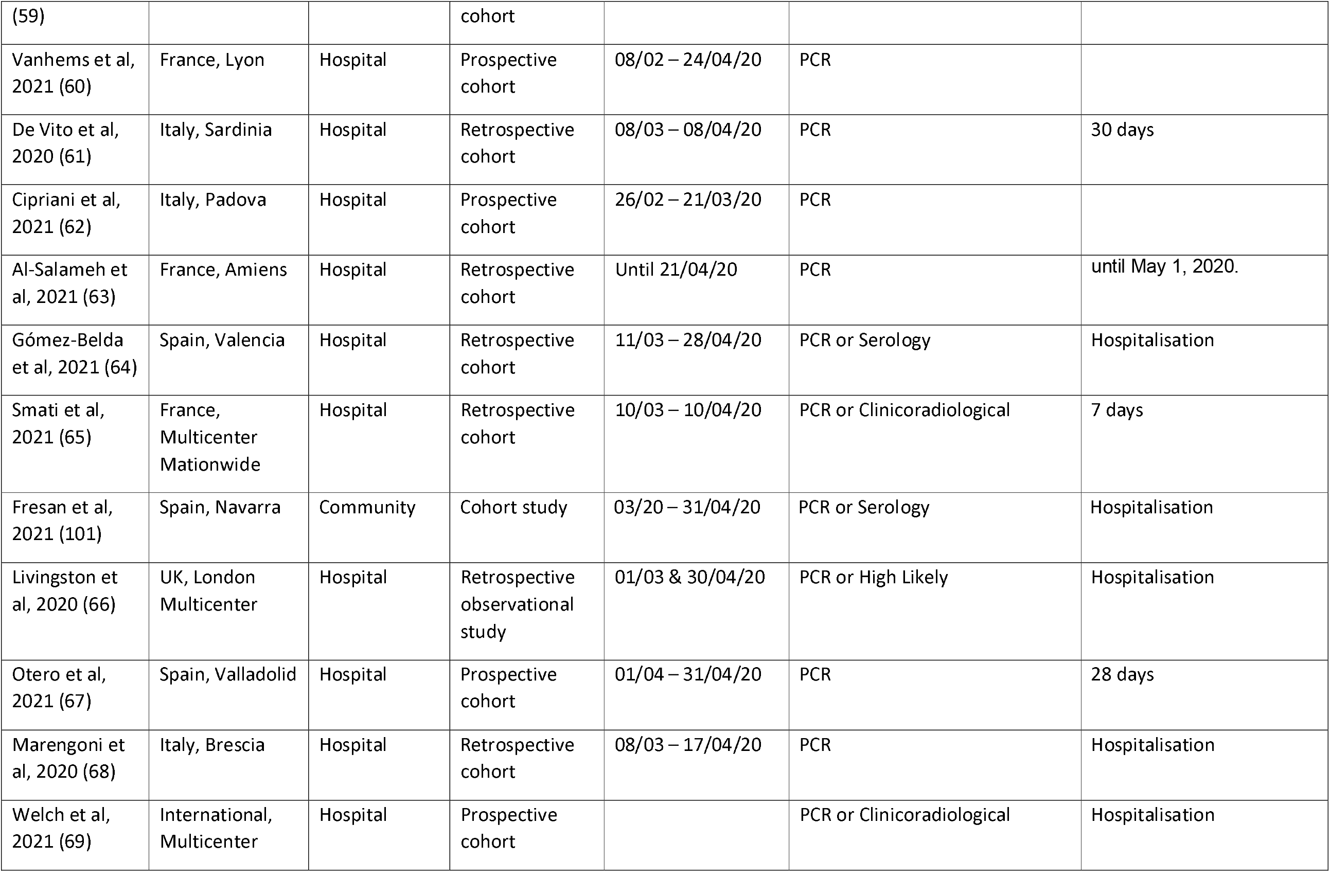
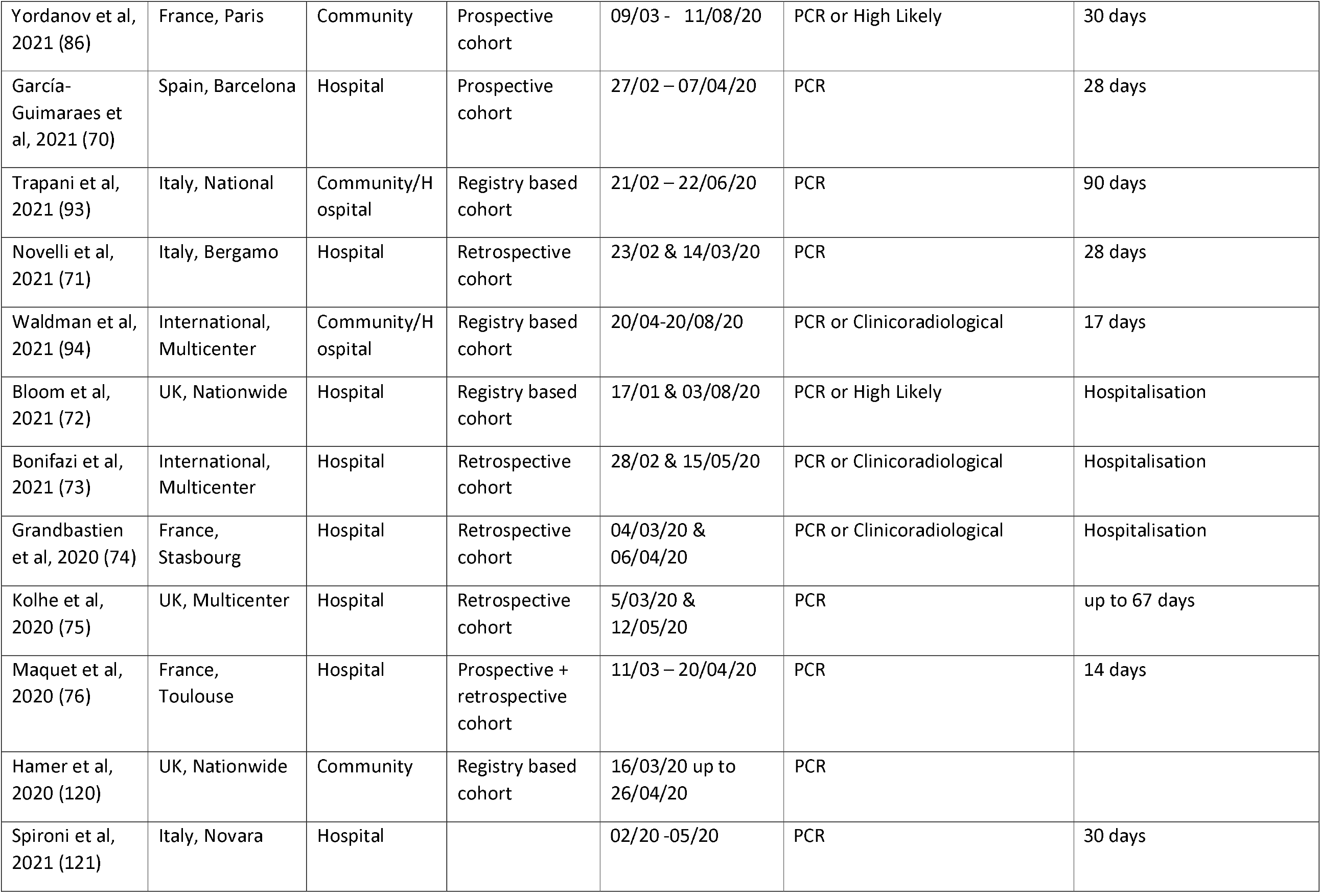
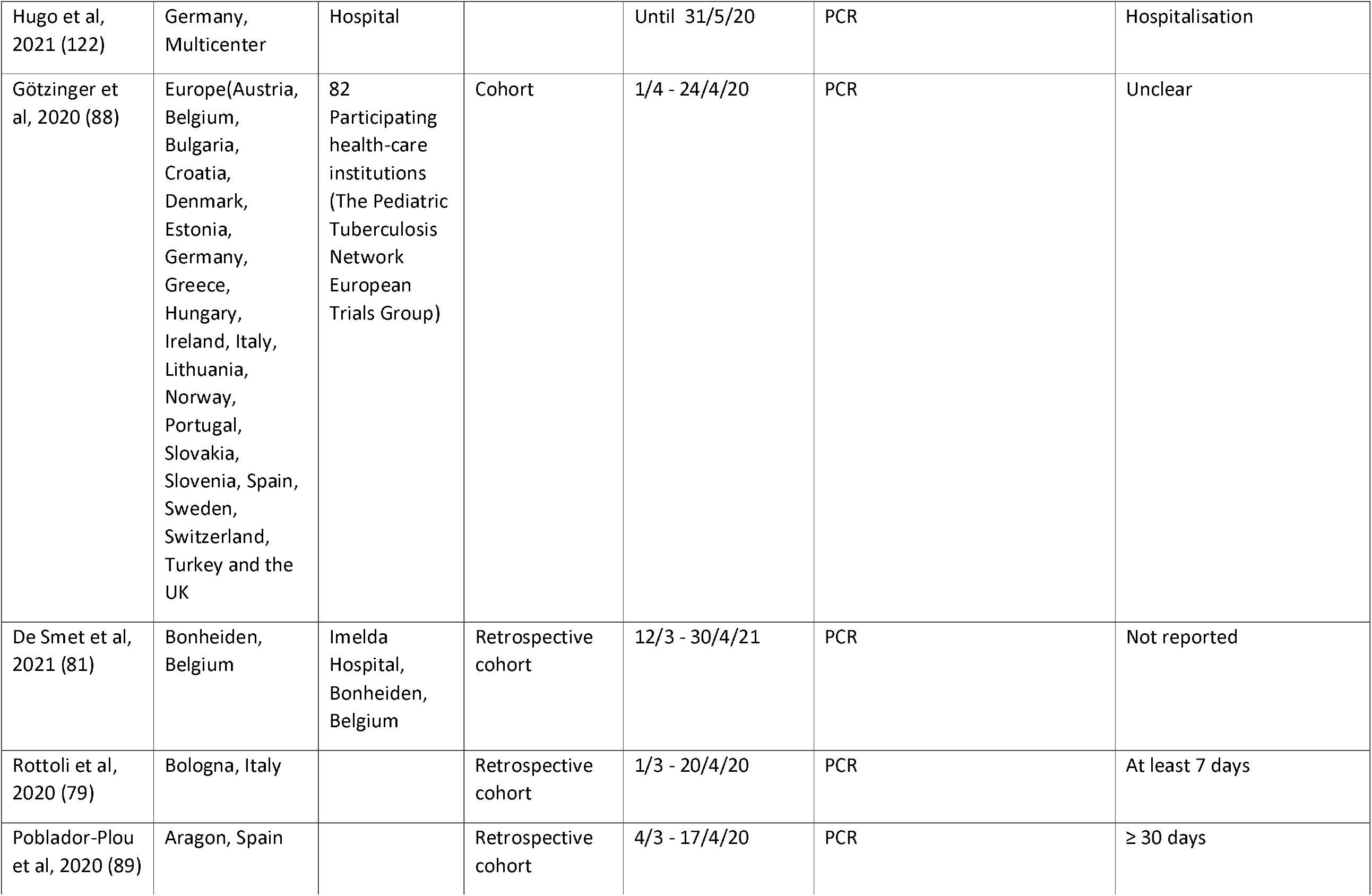
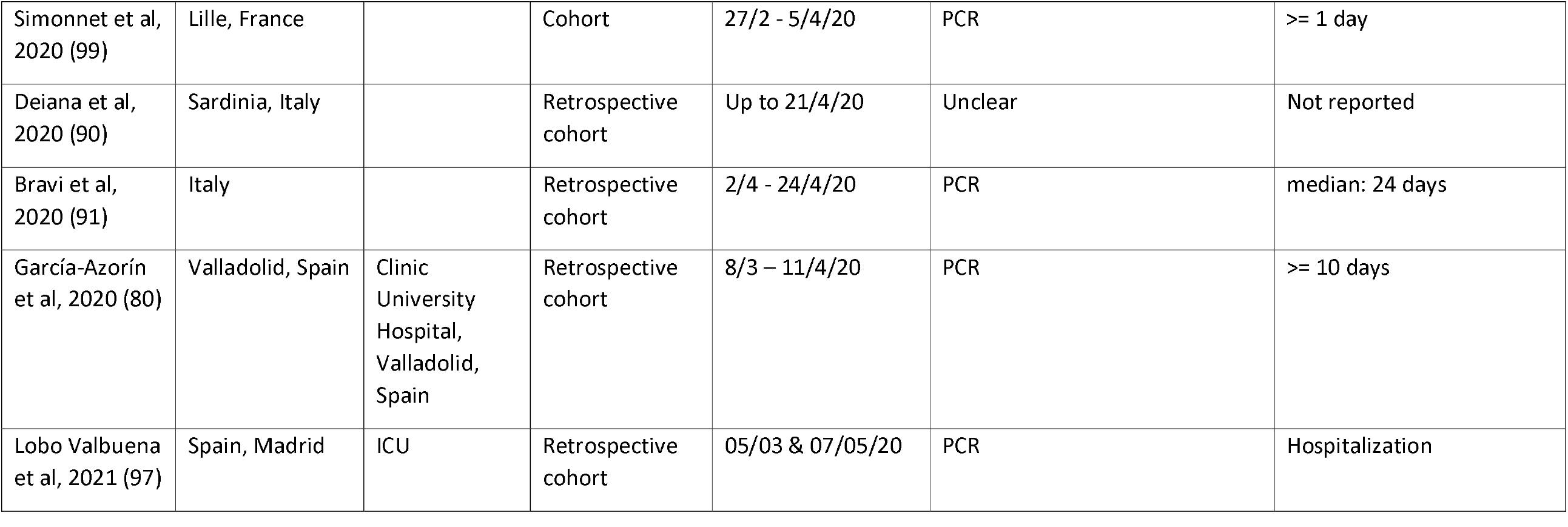

## Appendix 3. Risk of Bias and Quality appraisal

The JBI critical appraisal tool for cohort studies consists of eleven questions evaluating the appropriateness of the sample used in each study, the recruitment methods, the measurements of exposures and outcomes, the identification and managing strategies for confounding factors, the follow up process and the statistical analysis. For the purposes of this study, we accepted as accurate exposures (e.g., age, sex, comorbidities) and outcome measures documented by the treated clinician or captured from health databases. We considered age, gender and recruitment setting (a surrogate measure of baseline COVID-19 severity) the most important confounding factors. All the included studies clearly grouped patients based on the recruitment setting. Therefore, we scored high risk of bias for studies that were not at least age and gender adjusted (Q5). Moreover, since the aim of our study is to assess the risk factors for adverse COVID-19 outcomes among people who may contract the disease in the future, we considered that adjustment for presenting clinical characteristics associated with COVID-19 (e.g., symptoms, oxygen saturation etc) would be a source confounding in our study. Therefore, in such cases, we scored high risk of bias in the last domain of the instrument (Q11).

Studies that provided appropriately adjusted analyses for some but not all variables were judged to be at low risk of bias for confounding. However, we noted which variables were adjusted and conducted a sensitivity analysis only including adjusted variables, the results of which are presented in the main text of the manuscript. Finally, we considered 2 weeks to be adequate follow-up to capture the outcomes of interest, as the median length of hospital stay varied between 5-13 days within the included studies, and the median length of ICU admission between 7-9 days. Therefore, 2 weeks of follow-up would suffice to capture the outcomes of significantly more than half of the participants. Studies only including patients that were discharged or deceased were also judged to be at low risk of bias.

JBI critical appraisal tool for cohort studies

|  |  |
| --- | --- |
| Q1 | Were the two groups similar and recruited from the same population? |
| Q2 | Were the exposures measured similarly to assign people to both exposed and unexposed groups? |
| Q3 | Was the exposure measured in a valid and reliable way? |
| Q4 | Were confounding factors identified? |
| Q5 | Were strategies to deal with confounding factors stated? |
| Q6 | Were the groups/participants free of the outcome at the start of the study (or at the moment of exposure)? |
| Q7 | Were the outcomes measured in a valid and reliable way? |
| Q8 | Was the follow up time reported and sufficient to be long enough for outcomes to occur? |
| Q9 | Was follow up complete, and if not, were the reasons to loss to follow up described and explored? |
| Q10 | Were strategies to address incomplete follow up utilized? |
| Q11 | Was appropriate statistical analysis used? |

## Appendix 4

**Figure 4a-b.**
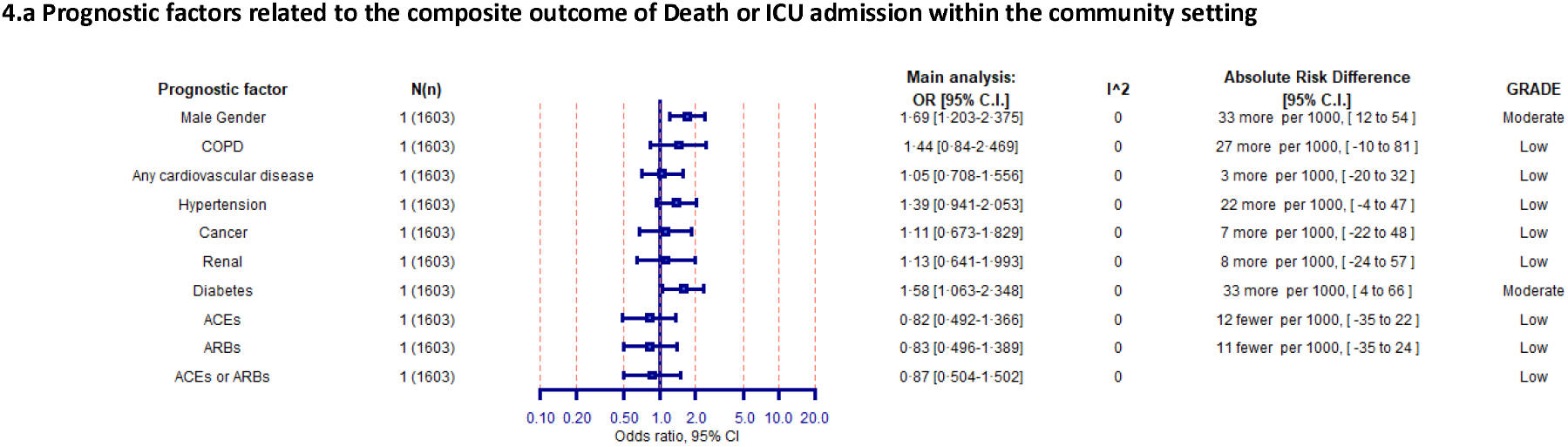
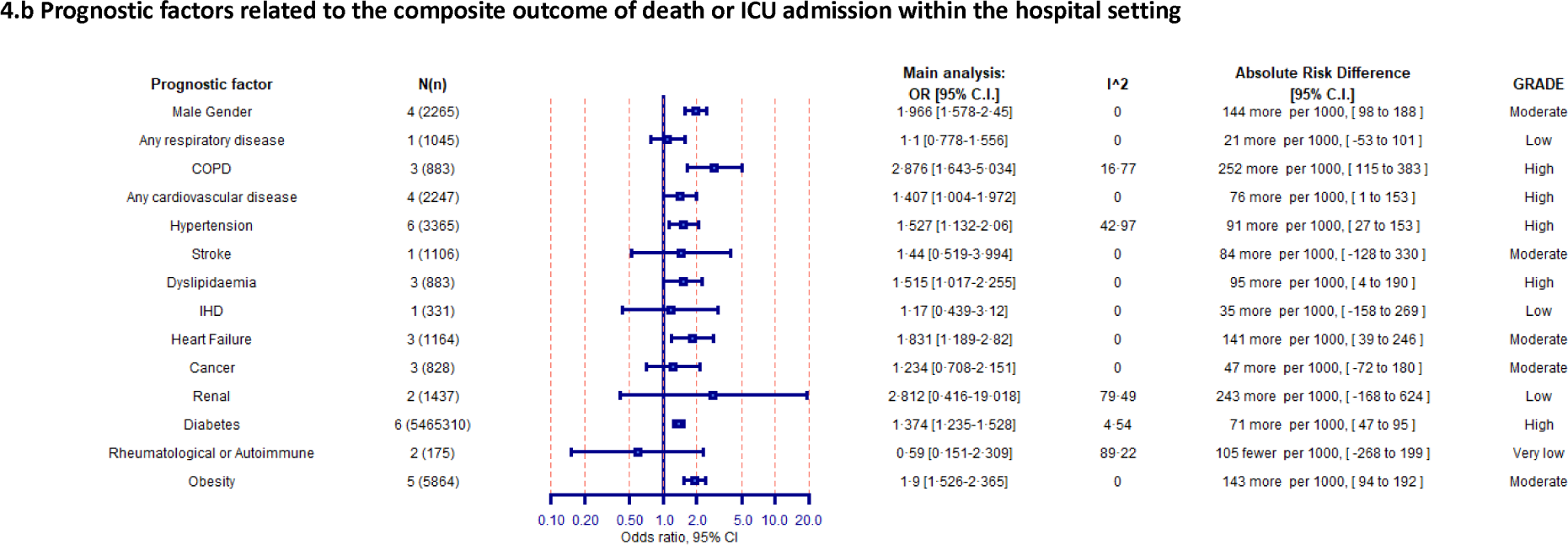
Forest plot showing the summary of available evidence regarding the association between prognostic factors and the composite outcome death or ICU admission due to COVID-19 within the community (a), and hospital setting (b).

## Appendix 5

**Figure 5a-b.**
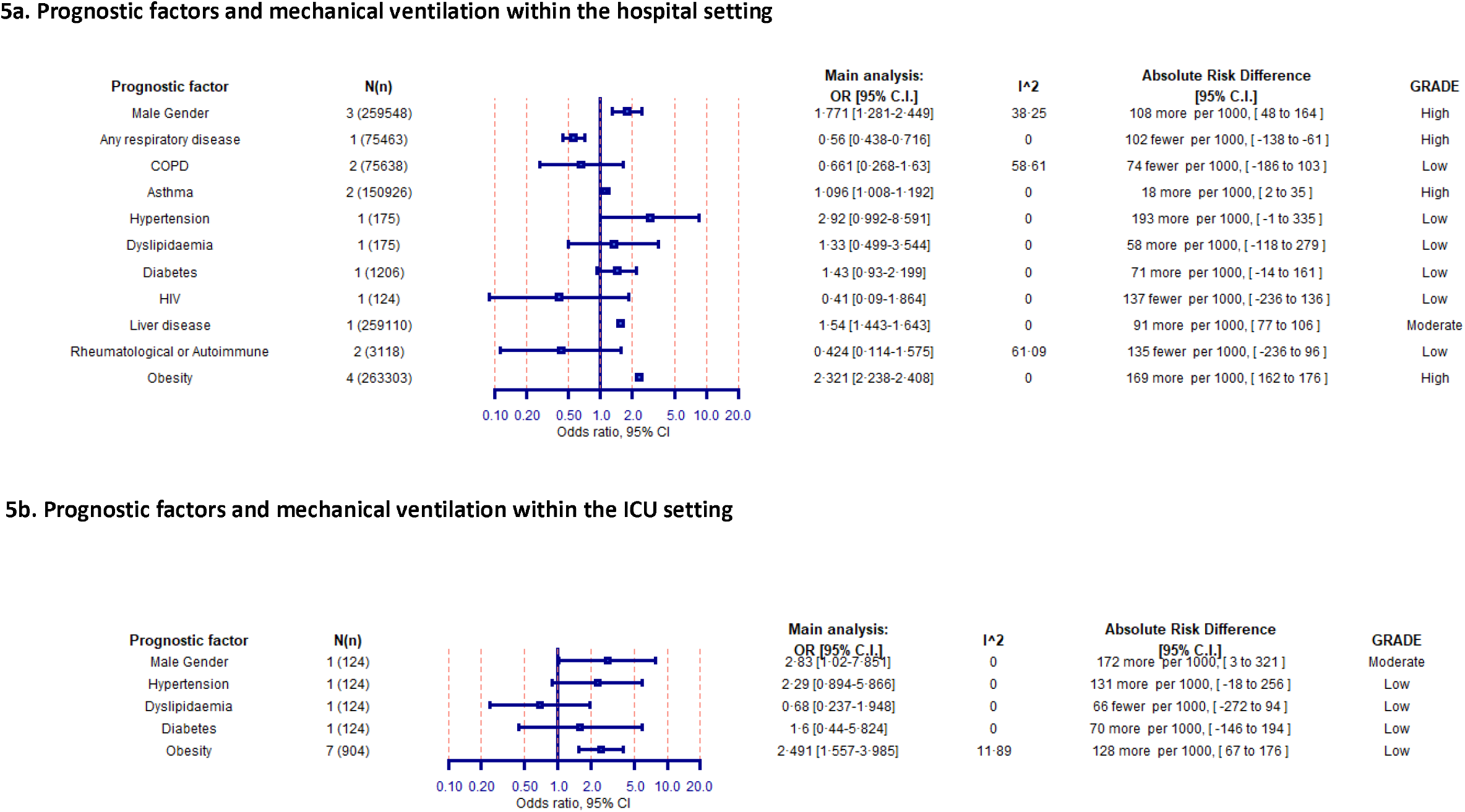
Forest plot showing the summary of available evidence regarding the association between prognostic factors for mechanical ventilation due to COVID-19 among those from within the hospital (6a) and ICU (6b) setting.

